# Patterns of air pressure, wind speed, and temperature are correlated with an increased risk of clinical infection from *Vibrio vulnificus* in endemic areas

**DOI:** 10.1101/2022.05.20.22275342

**Authors:** Andrea J. Ayala, Ketty Munyenyembe, Salvador Almagro-Moreno, C. Brandon Ogbunugafor

**Affiliations:** Department of Ecology and Evolutionary Biology, Yale University, New Haven, Connecticut, United States; Burnett School of Biomedical Sciences, University of Central Florida, Orlando, Florida, United States; National Center for Integrated Coastal Research, University of Central Florida, Orlando, Florida, United States; Public Health Modeling Unit, Yale School of Public Health, New Haven, Connecticut, United States

## Abstract

*Vibrio vulnificus* remains one of the deadliest waterborne pathogens, yet little is known of the ecology that drive outbreaks. As a nationally notifiable disease, all cases of *Vibrio vulnificus* diagnosed in the United States are reported to the state in which they occurred, as well as the Centers for Disease Control (CDC) in Atlanta, Georgia. Given that the state of Florida is a ‘hotspot’ for *Vibrio vulnificus* in the United States, we examined the prevalence and incidence of cases reported to the Florida Department of Health (2008-2020). Using a dataset comprised of 448 cases of disease caused by *Vibrio vulnificus* infection, we identified environmental variables that were associated with clinical cases and deaths. Combined with data from the National Oceanic and Atmospheric Administration (NOAA), we developed statistical models to examine the relationship between meteorological measurements such as wind speed, air temperature, water temperature, and sea-level pressure. We then examined the association of those meteorological variables with coastal cases of *Vibrio vulnificus*, including the outcome, survival, or death. Between 2008 and 2020, *Vibrio vulnificus* cases generally increased over time, peaking in 2017. Not surprisingly, there appears to be a strong correlation between water temperature and air temperature in Florida. However, as water temperature and air temperature increased, so too did the likelihood that an infection with *Vibrio vulnificus* would lead to patient death. Interestingly, we also found that as mean wind speed and sea-level pressure decreased, the probability that a *Vibrio vulnificus* case would be reported increased. Given these results, we discuss the potential factors that may contribute to the observed correlations. We further speculate that the meteorological variables we measured may increase in importance as they relate to the incidence of *Vibrio vulnificus* in light of rising global temperatures.

## INTRODUCTION

Facultative pathogens are free-living microorganisms that are characterized by their ability to persist, reproduce, and transmit to susceptible hosts directly from their natural habitats [1]. Many are the causative agents of significant world-wide public health burdens, and yet, given their environmental sourcing, cannot be eradicated [2, 3]. Some members of the Vibrionaceae, a family of aquatic bacteria, rank among the best known of facultative pathogens, with broad niche breadths and a nearly pan-coastal distribution [4]. Classified within the Vibrionaceae is *Vibrio vulnificus*, one of the most virulent *Vibrio* species [5]. The bacterium is a halophilic, autochthonous inhabitant of aquatic environments that causes a life-threatening septicemia in susceptible hosts, with a reported case fatality rate of up to 50 percent [5]. Although *V. vulnificus* is known for its worldwide distribution, epidemiological reports are most commonly linked to temperate coastal nations such as the United States, France, Germany, Israel, Korea, and Taiwan [6, 7, 8].

Living in Florida, and other states on the Gulf Coast (Texas, Louisiana, Mississippi, Alabama) is associated with an increased risk of contracting *V. vulnificus*, primarily between May and October [9, 10, 11]. *V. vulnificus* is also often isolated from locations where salinity ranges from 15 to 25 parts per thousand (ppt), and the water temperatures range from 9° to 31°C [10, 12, 13, 14]. In general, prior studies performed in Florida have predominantly assessed the clinical features of infection as opposed to analyzing the ecological patterns that occur in association with case reports [5, 15, 16, 17, 18]. However, there are studies that previously examined the relationship between *V. vulnificus* clinical cases and temperature [19, 20]. Even before mandatory CDC reporting, cases were detected consistently between March and November, with a peak period of incidence in May [5, 15]. Due to its high mortality rate, *V. vulnificus* was declared the most lethal foodborne illness in the state in 1993 [15]. Since then, the Florida Department of Health has maintained a reporting database, which has demonstrated an increased incidence of *V. vulnificus* clinical cases [16].

In addition to these studies of clinical infection, there are several examinations of the relationship between *V. vulnificus* abundance and temperature or seasonality. A study of four coastal habitats found that elevated sea surface temperatures and suspended particulate matter were variables that were predictive for detecting *V. vulnificus* both in water samples and oysters [21]. López-Pérez et al. (2021) reported recovering *V. vulnificus* in Florida more often during months when the water temperature exceeded 20°C [22]. With respect to seasonality, significant fluctuations were detected in the abundance of *V. vulnificus* in several aquatic organisms across several Gulf states [23, 24]. Thus, we cannot overstate the importance of water and ambient air temperature to the ecology of *V. vulnificus* [22, 25]. That said, there remain many other abiotic factors which may contribute to *V. vulnificus* emergence that may contribute to poor clinical outcomes.

In this study, we utilize data science approaches to establish associations between clinical cases of *V. vulnificus* and standardized meteorological conditions: water temperature, wind speed, air temperature, and sea-level pressure during months were case reported. Using Florida as a model setting, we use a combination of case data from the Florida Department of Health in conjunction with data from the National Oceanic and Atmospheric Administration (NOAA) to identify several meaningful correlations. First, we examined correlations between our four variables of interest: wind speed, sea-level pressure, water temperature, and air temperature. Among these, we found that only air temperature and water temperature were strongly correlated. We next identified several significant associations between ecological variables and risk of infection. Specifically, as wind speed and sea-level pressure decreased, the likelihood of patient death increased. On the other hand, as air and water temperature decreased, so did the prospect of surviving an infection with *V. vulnificus*. In closing, we speculate on the mechanisms underlying these associations and reflect on their relevance for conversations surrounding climate change and epidemiology.

## MATERIALS AND METHODS

### Terminology

In this study, we use the term “environmental variables” to describe the various factors that were used to diagnose correlations with clinical cases. The data themselves come from meteorological measurements and are key features of the microbial ecology that underlie *V. vulnificus* infections.

### Case acquisition and Case Fatality Rates

Redacted data on *V. vulnificus* cases were kindly provided to the Ogbunu Lab at Yale University in December 2020 by the Florida Department of Health [16]. Pathogenic *Vibrio* illnesses became notifiable diseases to the Cholera and Other Vibrio Illness Surveillance (COVIS) system and Foodnet in late 2007. Since then, the state of Florida documented 448 cases of *V. vulnificus* between 1/1/2008 and 12/20/2020 that were acquired in the state and traceable to their originating county. Outcomes for each case, e.g., survival or death, were also provided in the dataset. To calculate the case fatality rate (CFR) from our 448 cases, we divided the total number of deaths, e.g., 111 that occurred between 2008 and 2020 by 448. We then multiplied the resulting ratio by 100 to yield a percentage.

### Clinical Cases and Meteorological Data

In order to correspond meteorological information with our county case data, we utilized publicly available meteorological information from the global National Oceanic and Atmospheric Administration (NOAA) Marine Environmental Buoy Database and the NOAA National Buoy Data Center (NBDC) [26, 27] (Fig. 1). Florida’s NOAA buoys are distributed between two systems, the Gulf of Mexico-(West) and the Gulf of Mexico-(East), across a coastline length of 2,170 km. Each coastal buoy, regardless of system, collects standard meteorological data on a per-six minute interval basis for the following variables: year, month, date, hour, minute, wind, wind speed (WSPD) in meters/second, gust, wave height in meters, waves, waves at the dominant period, average wave period in seconds, direction of the waves at the dominant period, sea-level pressure in hectopascals (PRES), ambient air temperature in degrees Celsius (ATMP), water temperature in degrees Celsius (WTMP), dewpoint temperature in degrees Celsius, station visibility in nautical miles, pressure tendency and tide, e.g., the water level in feet above or below mean lower low water. Historical data is available for most NOAA buoys dating back to January 1^*st*^, 2005.

**Figure 1.**
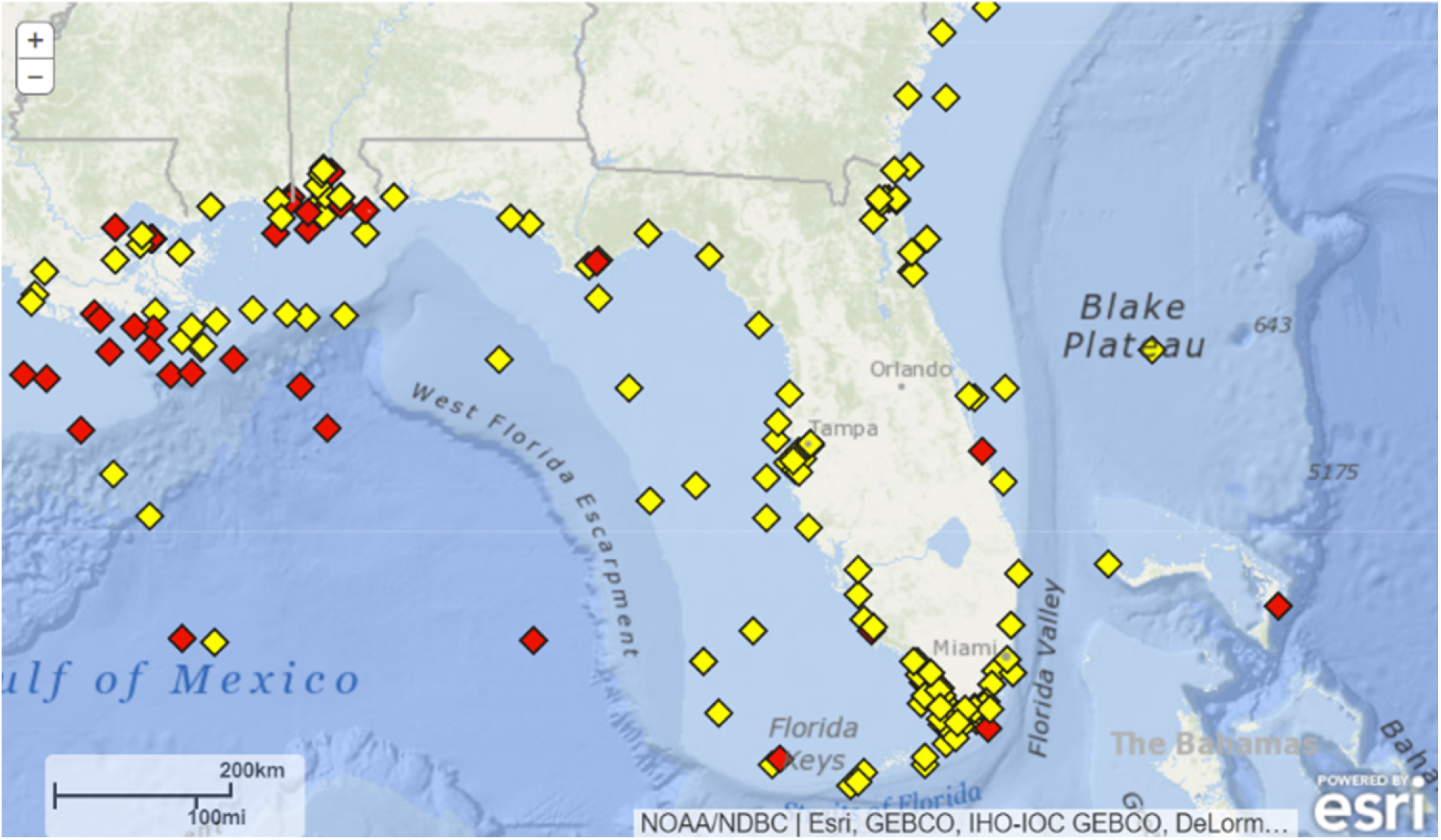
NOAA Coastal Buoys of Florida. Reprinted from https://www.ndbc.noaa.gov/ on 12/22/2021. This demonstrates the distribution of the NOAA coastal buoys (in yellow and red) around the coast of Florida, which collect ecological and environmental data on coastal conditions every six minutes. Buoys in yellow are up to date on data collection, whereas buoys in red have not transmitted data within the last 24 hours since the image was downloaded.

We corresponded NOAA buoys (Fig. 1) to Florida coastal counties first by calculating which NOAA buoys were the closest to the middle of the coastline of the county, according to the centroid. Secondly, from this information, we identified which of the closest buoys maintained historical meteorological data dating back to 2008, the year that V. vulnificus cases were consistently reported to COVIS and FoodNet. Due to uneven coverage of the Florida coast by individual NOAA buoys, multiple coastal counties were often assigned a single buoy. Historical data (2008 – 2020) was extracted from NOAA, according to the corresponding buoy (Table 1), using the R programming language. Subsequently, the data was downloaded as txt files from each NOAA buoy’s page into the R statistical language platform for windspeed (WSPD), atmospheric temperature (ATMP), water temperature (WTMP) and sea-level pressure (PRES). Missing buoy data were indicated as 99 (WSPD), 999 (ATMP), 999 (WTMP) and 9999 (PRES) by NOAA. These values were removed from the imported data and means were calculated for all months (January to December) for each county, for the years of interest, 2008 to 2020. Results were saved as csv files and the data were transposed in excel with 1-13 corresponding to 2008-2020 and 1:12 corresponding to January-December. All data was then uploaded into publicly available google sheets for downstream statistical analyses.

**Table 1.**
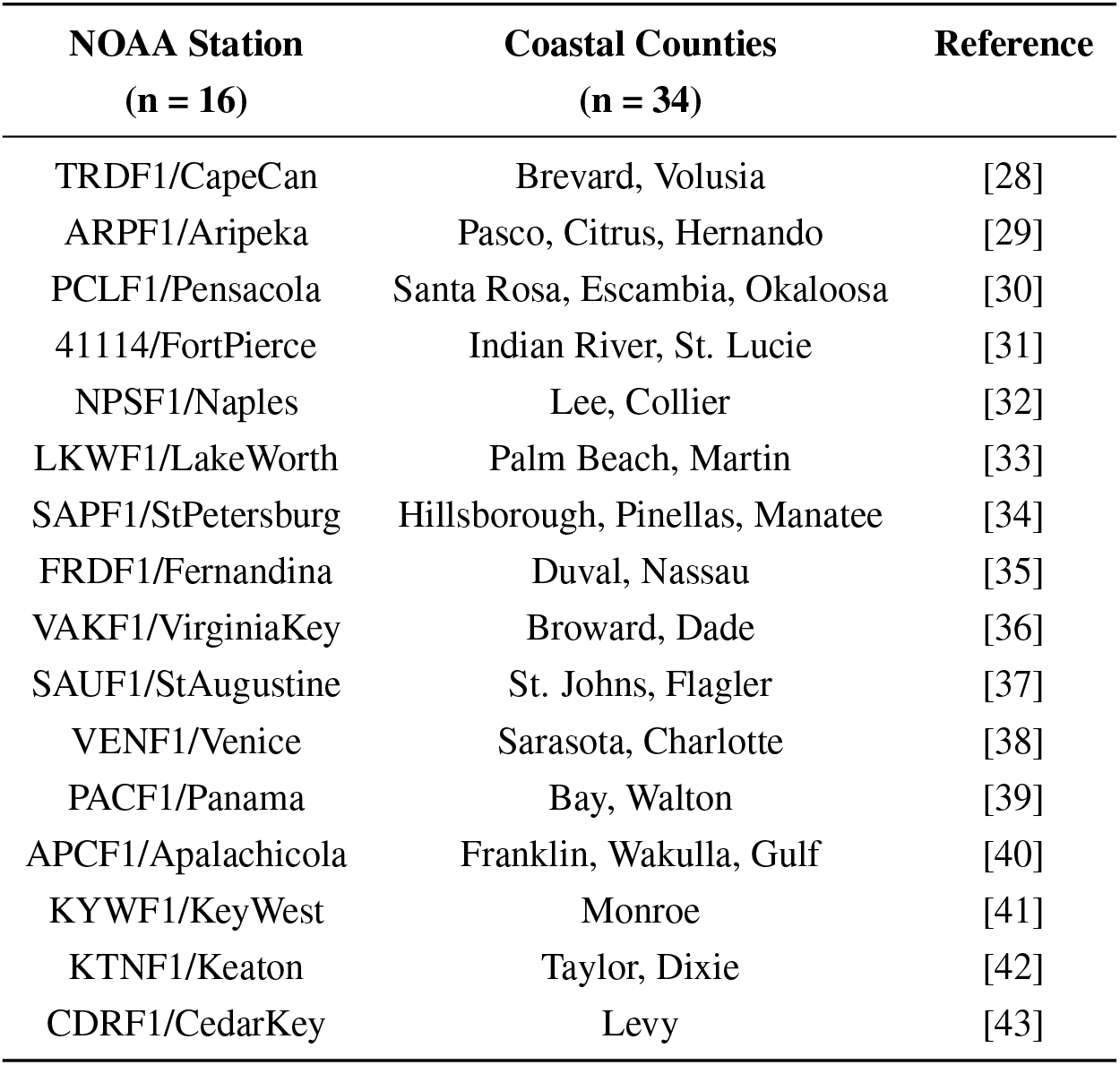
Names of NOAA buoys used in our analysis. The Florida counties that correspond to meteorological data, and NOAA reference information for each buoy.

Data was only analyzed from each NOAA buoy for the year that a case occurred in its corresponding county. For the remainder of the case year, during months that no cases were reported, we also collected the monthly mean of the listed variables. This provided the opportunity to compare the meteorological conditions that occurred during months when cases were reported, and during months that cases were not reported.

Inclusion criteria for environmental variables. Note that out of all the data collected by the NOAA buoys, we only extracted and analyzed correlations with wind speed (WSPD), air temperature (ATMP), water temperature (WTMP) and sea-level pressure (PRES). As first proposed in the introduction, these are related to factors associated with *Vibrio* populations in other contexts. Rather than analyze correlations between all available NOAA environment variables, our approach was more directed and hypothesis-driven, utilizing only those meteorological measurements that we believe might have a relationship with *Vibrio* populations [22]. Below we describe the variables.

- *Storm events* (WSPD, PRES): Tropical cyclones have been linked to a rise in *V. vulnificus* cases [44, 45, 46, 47]. After Hurricane Katrina made landfall in Louisiana, extensive flooding in the city of New Orleans was linked to a spike in *V. vulnificus* clinical cases [44]. A longitudinal study of Lake Pontchartrain post-Hurricanes Katrina and Rita in New Orleans found that *V. vulnificus* abundance increased closer to the lake shoreline, and was positively correlated to increasing temperature, turbidity, and salinity [46]. Interestingly, sampling in the Chesapeake Bay area of Maryland, USA, found no significant change in the concentration of *V. vulnificus* in samples collected from the top of the water column and in oysters before and after Hurricane Irene impacted the area, likely as a result of the ensuing wave action and sediment resuspension [48, 49]. In North Carolina, data on the abundance of *V. vulnificus* within the Neuse River Estuary was collected before and after two major storm events – Hurricane Ophelia and Tropical Storm Ernesto [45]. Unexpectedly, the prevalence of *V. vulnificus* in storm samples varied, from Tropical Storm Ernesto, with an 80% prevalence in storm samples, to Hurricane Ophelia, with a 31% storm sample prevalence [45].
- *Elevated temperature* (WTMP, and ATMP). Water temperature has an established relationship with *V. vulnificus* density and abundance. Elevated water temperature has the highest association with *V. vulnificus* abundance, while other variables, such as increased turbidity, low dissolved oxygen concentrations, increased estuarine bacterial levels and high fecal coliform levels are also predictive for the detection of *V. vulnificus* [50]. Though most of the literature has focused on water temperature, we wanted to test if there was a relationship with air temperature.
- *Seasonality*. In the northeastern United States, Tilton and Ryan (1987) sampled water and shellfish, specifically mussels, oysters, clams, and whelk, for the presence and abundance of *V. vulnificus* during the spring and summer months [19]. No *V. vulnificus* could be isolated from water samples and shellfish until the temperature reached 17°C, plateauing when temperatures reached 22°C [19].

### Statistical Analyses

All statistical analyses were performed using the R statistical language (version 4.1.2) [41]. We assessed the normality of our data distributions for Seasonality, WSPD, PRES, ATMP, and WTMP using the Shapiro-Wilks test of Normality, with a p-value threshold of 0.05 [42]. To determine the strength of the relationship between each meteorological variable, we performed a series of non-parametric Spearman’s Rank Correlation *ρ* coefficients to measure their association. As our meteorological variables are in a continuous data format, Spearman’s Rank Correlation *ρ* is the most appropriate test to measure associations between the data. We performed the analyses covering four conditions: all periods, e.g., during months when no cases were reported, during months when cases were reported, and during months when deaths were reported as one dataset. Then, we subsetted the data for ‘all periods’ to examine the strength of those relationships during each condition: months when no cases were reported, months when cases were reported, and months when deaths were reported. Subsequent analyses of stratified data were performed using the non-parametric Kruskal-Wallis H test for three independent groups [43], and the Mann-Whitney U test for two independent groups [44], both with a p-value threshold of 0.05 for statistical significance.

Associations between our meteorological variables and seasonality were assessed using the non-parametric Chi-Square Goodness of Fit test. Although our meteorological variables are in a continuous data format, seasonality is represented by categorical data, e.g., winter, spring, summer, and fall. This statistical test allowed us to examine whether a given phenomenon, i.e., seasonality of the reported cases, adhered to a proposed distribution of the data. Specifically, we asked whether each season deviated from a distribution where cases were distributed across seasons equally.

To determine the most meaningful model overall model, we ran a series of logistic regressions using combinations of our environmental variables coupled with County, NOAA reporting stations and their corresponding counties, as well as Month, and Season as our variables of interest. Logistic regression was utilized by coding months when cases occurred as one, and months when cases did not occur as zero [28]. We used AIC and McFadden’s pseudo *R*^2^ to identify the model that accounts for the predominance of the variation seen in our data [45, 46].

## RESULTS

### Descriptive Analysis of Coastal and Non-Coastal Cases Combined: Seasonality, County Occurrence, Annual Reporting, and Monthly Analysis

A total of 456 cases were reported between 1/1/2008 and 12/20/2020 to the Florida Department of Health. Eight cases were unconfirmed with respect to having been acquired in Florida, and thus were removed from the dataset, leaving 448 cases. Out of 67 total counties, *V. vulnificus* cases were reported in 50 (Fig. 2A). Counties reporting equal to or greater than 25 cases over the twelve-year period were: Hillsborough, Escambia, Brevard, Broward, and Pinellas. Counties reporting the fewest cases (one case each) were: Dixie, Franklin, Gadsden, Madison, Osceola and Putnam.

**Figure 2.**
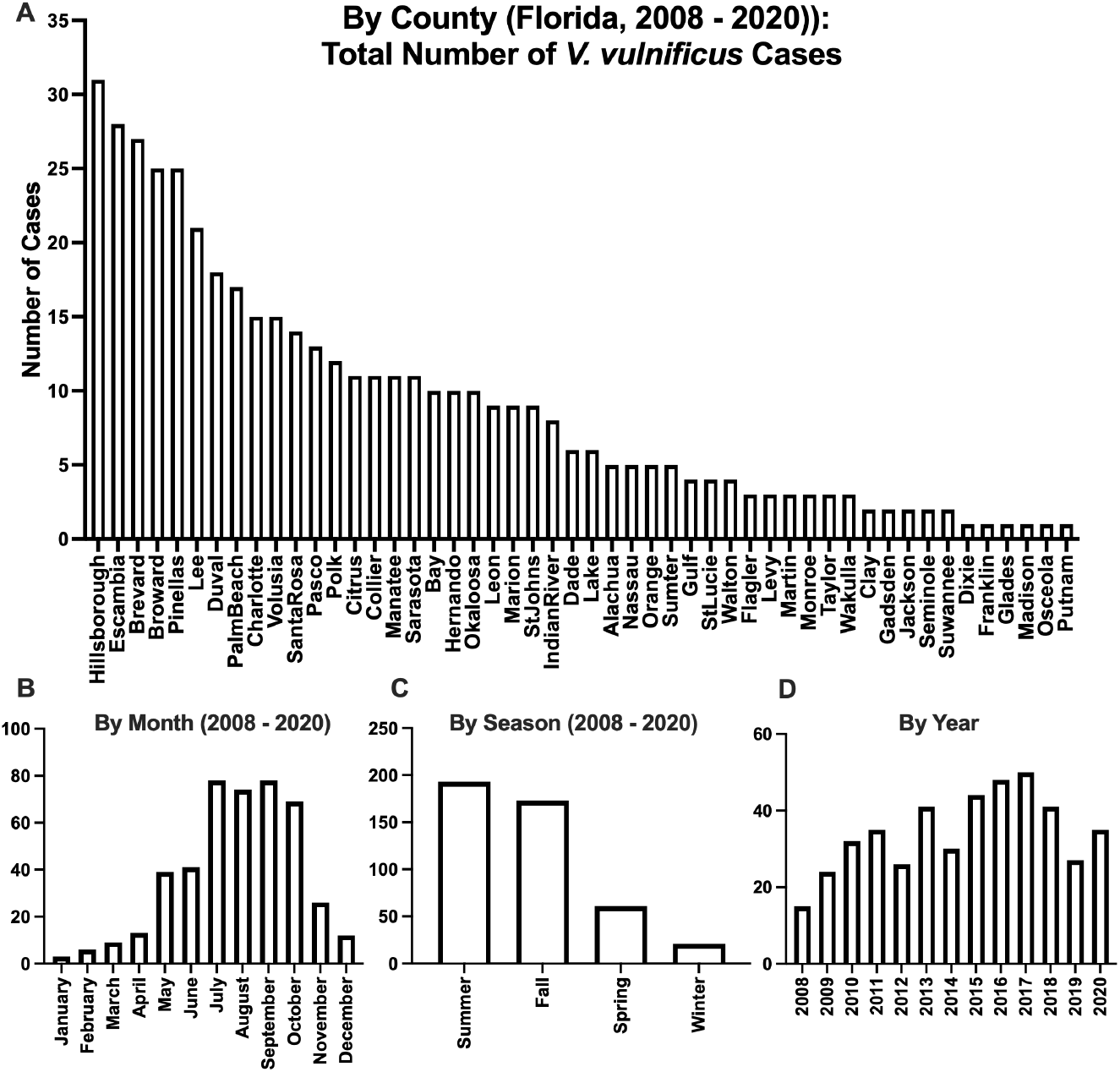
Occurrences of V. vulnificus in Florida between 2008 and 2020. Panel A depicts the relative occurrences of the 448 cases across each of Florida’s 50 reporting counties, that were documented between 2008 and 2020. The data is in descending order of frequency by county, with the county of Hillsborough demonstrating the highest reported occurrences of V. vulnificus in the state. Panel B demonstrates the breakdown of V. vulnificus cases in Florida by month, between the years of 2008 and 2020 in the state of Florida. July, August, and September demonstrate the highest incidence of reporting. Panel C breaks down our dataset of 448 cases according to season. Summer (June, July, and August) had the greatest number of reported cases, followed by fall (September, October, and November), with the fewest cases reported in the winter (December, January, and February), followed by spring (March, April and May). Panel D represents our dataset of 448 cases broken down according to year. In 2008, the fewest cases were reported, while in 2017, the highest frequency of cases was reported.

We also found that cases were reported most frequently during the months of July and September (Fig. 2B) during the period spanning from 2008 and 2020, with the fewest cases occurring in January and February. Seasonality as a variable was also assessed (Fig. 2C), with the most cases reported in the summer months (June, July, and August) and the fewest in the winter (December, January, and February). The Shapiro-Wilk’s Test of Normality demonstrated that the data for seasonality was normally distributed (W = 0.88475, p-value = 0.3593). Nonetheless, we utilized a non-parametric Chi-Square Goodness of fit test, given our question, “are cases statistically distributed across seasons unequally?” Our sample sizes for each season were: Winter, n = 21, Spring, n = 61, Summer, n = 193, and Fall, n = 173. This led to a significant Chi-Square *χ*^2^ = 3, df = 3, p-value = <0.0001, indicating that seasonality was statistically significant. We followed up this omnibus Chi-Square Goodness of Fit test with sequential Chi-Square Goodness of Fit Tests, corrected using a Bonferroni correction adjusted to a p-value of 0.00833. Winter-spring was significantly reported as *χ*^2^ = 19.512, df = 1, p-value = <0.0001, winter-summer was significantly reported as *χ*^2^ = 119.09, df = 1, p-value = <0.0001, winter-fall was significantly reported as *χ*^2^ = 138.24, df = 1, p-value = <0.0001, spring-summer was significantly reported as *χ*^2^ = 54.607, df = 1, p-value = <0.0001, spring-fall was significantly reported as *χ*^2^ = 68.598, df = 1, p-value = <0.0001, and summer-fall was not significant at *χ*^2^ = 1.0929, df = 1, p-value = 0.2958.

When trends were separated by year (Fig. 2D), they demonstrate that the fewest cases were reported in 2008, and the largest number in 2017. In general, cases increased on an annual basis. Specifically, in 2008, n = 15 cases were reported, and in 2017, n = 50 cases were reported, for an increase of 333% between the lowest and highest reporting yearly incidence in Florida.

### Overall Case Fatality Rate and Mortality and Case Fatality Rate by Month, Season, and Year

We calculated the overall case fatality rate (CFR) from our 448 cases, of which 111 cases had death as an outcome. This yielded a CFR of 24.78% across the state of Florida in our given time period [47]. When categorized by month, we found that like our case occurrence data, the overall deadliest months were September (21 deaths), followed by July (19 deaths). The fewest deaths occurred in February (0 deaths), which were followed by January and March (2 deaths each). CFRs by month are given in Table 2.

**Table 2.** Case fatality rates of *V. vulnificus* as reported by the state of Florida. In general, case fatality rates did not exceed 30 percent, except for the months of January and April.

| Month | Case Fatality Rate |
| --- | --- |
| January | 66.67% |
| February | 0 |
| March | 22.22% |
| April | 30.77% |
| May | 28.21% |
| June | 24.39% |
| July | 24.36% |
| August | 24.32% |
| September | 26.92% |
| October | 21.74% |
| November | 26.92% |
| December | 16.67% |

Our computed CFRs suggested that cases were more likely to have a negative outcome from infection during the months of December, followed by April. When categorized by season, the data showed that the case fatality rate of the fall was 24.86%, the CFR of the spring was 27.87%, the CFR of the summer was 24.35%, and the CFR of the winter was 19.05%. Yearly CFRs were as follows: 2008 (40%), 2009 (29.17%), 2010 (31.25%), 2011 (34.28%), 2012 (34.62%), 2013 (29.27%), 2014 (16.67%), 2015 (29.55%), 2016 (20.83%), 2017 (22%), 2018 (17.07%), 2019 (7.4%), and 2020 (20%).

### Non-parametric Spearman’s Rank Correlation Tests

We found that many variables had statistically significant relationships according to a p-value of 0.05, across all stratifications, i.e., all data points, then subsetted across cases, no cases, and deaths (Table 3). However, the strengths of those correlations were weak except for ATMP and WTMP, where a strong relationship was defined as one with a *ρ* of greater than 80. Notably, WSPD was statistically significant and had a higher *ρ* correlation value with ATMP and WTMP during months when cases and deaths were reported, as opposed to when no cases were reported. Given the negative values attributed to the *ρ* values overall, this suggests that as WSPD increased, WTMP and ATMP decreased during months when cases and deaths were reported. In addition, WSPD and PRES were statistically significant during months when no cases were reported.

**Table 3.** Spearman’s Correlation Coefficients for Our Meteorological Variables. To determine the strength of the relationship between each meteorological variable, we performed a series of non-parametric Spearman’s Rank Correlation *ρ* coefficients to measure their association. We performed the analyses covering four conditions: all periods, e.g., during months when no cases were reported, during months when cases were reported, and during months when deaths were reported as one dataset. While many of the variables had statistically significant relationships according to the p-value, those correlations’ strengths were weak, except for ATMP and WTMP, where a strong relationship was defined as one with a *ρ* of greater than 80.

| Condition | Variable 1 | Variable 2 | S | p-value | Spearman's Rank Correlation $\rho$ |
| --- | --- | --- | --- | --- | --- |
| All | WSPD | ATMP | 1075558076 | 0.0004096 | -0.08290868 |
| All | WSPD | WTMP | 1047396443 | 0.02018 | -0.05455459 |
| All | WSPD | PRES | 1047395100 | 0.02018 | -0.05455323 |
| All | ATMP | WTMP | 41404884 | <0.0001 | 0.9583121 |
| All | ATMP | PRES | 1635063628 | <0.0001 | -0.646238 |
| All | WTMP | PRES | 1637838280 | <0.0001 | -0.6490316 |
| No cases | WSPD | ATMP | 645444236 | 0.1158 | -0.03995624 |
| No cases | WSPD | WTMP | 6.26E+08 | 0.7344 | -0.008625618 |
| No cases | WSPD | PRES | 662269930 | 0.00826 | -0.06706622 |
| No cases | ATMP | WTMP | 25200928 | <0.0001 | 0.9593956 |
| No cases | ATMP | PRES | 1029502253 | <0.0001 | -0.6587603 |
| No cases | WTMP | PRES | 1030097863 | <0.0001 | -0.65972 |
| Cases | WSPD | ATMP | 3778459 | <0.0001 | -0.2462496 |
| Cases | WSPD | WTMP | 3701511 | 0.0003068 | -0.2208699 |
| Cases | WSPD | PRES | 3222070 | 0.3108 | -0.06273555 |
| Cases | ATMP | WTMP | 325697 | <0.0001 | 0.8925753 |
| Cases | ATMP | PRES | 4064933 | <0.0001 | -0.3407373 |
| Cases | WTMP | PRES | 4092443 | <0.0001 | -0.3498109 |
| Deaths | WSPD | ATMP | 34962 | 0.05402 | -0.2612656 |
| Deaths | WSPD | WTMP | 35597 | 0.03549 | -0.2841782 |
| Deaths | WSPD | PRES | 27074 | 0.866 | 0.02328938 |
| Deaths | ATMP | WTMP | 4681.5 | <0.0001 | 0.8311145 |
| Deaths | ATMP | PRES | 35431 | 0.03973 | -0.2781897 |
| Deaths | WTMP | PRES | 32235 | 0.2348 | -0.1628633 |

### Meteorological Data in Correlation with Case and Mortality Reporting

Given that NOAA buoys are only located along coastal regions, we analyzed how data from the buoys corresponded to cases that originated only in coastal counties, leaving a total of 383 cases for analysis, representing 34 counties (Table 1). When accounting for multiple counties and missing data, there were 1,813 total months during years when cases were reported for which we had meteorological data. A total of 263 of those months were months when cases were reported (n = 263). However, when accounting for multiple counties and missing data, there were 1,550 total months during years when cases were not reported. We then stratified WSPD (wind speed), ATMP (ambient air temperature), WTMP (water temperature) and PRES (sea-level pressure) across all coastal counties according to the months in a case year when no cases were reported, during months when cases with a survival outcome were reported, and during months when cases with a fatal outcome were reported for the following analyses.

#### a) Wind speed (WSPD)

The mean WSPD for months when no cases were reported was 3.35 ± 0.02 SE m/s (median = 3.24 m/s), while the mean WSPD for months when cases where survival was the outcome was a mean of 3.26 ± 0.05 SE m/s (median = 3.12 m/s), and the mean WSPD for months when death was the outcome was 3.27 ± 0.10 SE m/s (median = 3.04 m/s). Using the Shapiro-Wilks test of Normality, we found that WSPD deviated from a Gaussian distribution for all stratifications (W = 0.96096, p-value < 0.0001) when no cases were reported, (W = 0.89122, p-value = <0.0001) when survival was reported, and (W = 0.89771, p-value = <0.0001) when death was reported. Given that there was little difference between the means of months of V. vulnificus cases when deaths or survival were reported, we pooled our case data to perform a non-parametric Mann-Whitney U test to compare the means between the WSPD of the months when cases were reported, and months when cases were not reported. The Mann-Whitney U test indicated that the differences between the two groups were statistically significant (W = 325379, p-value = 0.01527), with the mean WSPD increased during the months that no cases were reported in contrast to months when cases were reported, regardless of their outcome (Fig. 3A). This suggests that as WSPD increases, the number of cases reported decreases.

**Figure 3.**
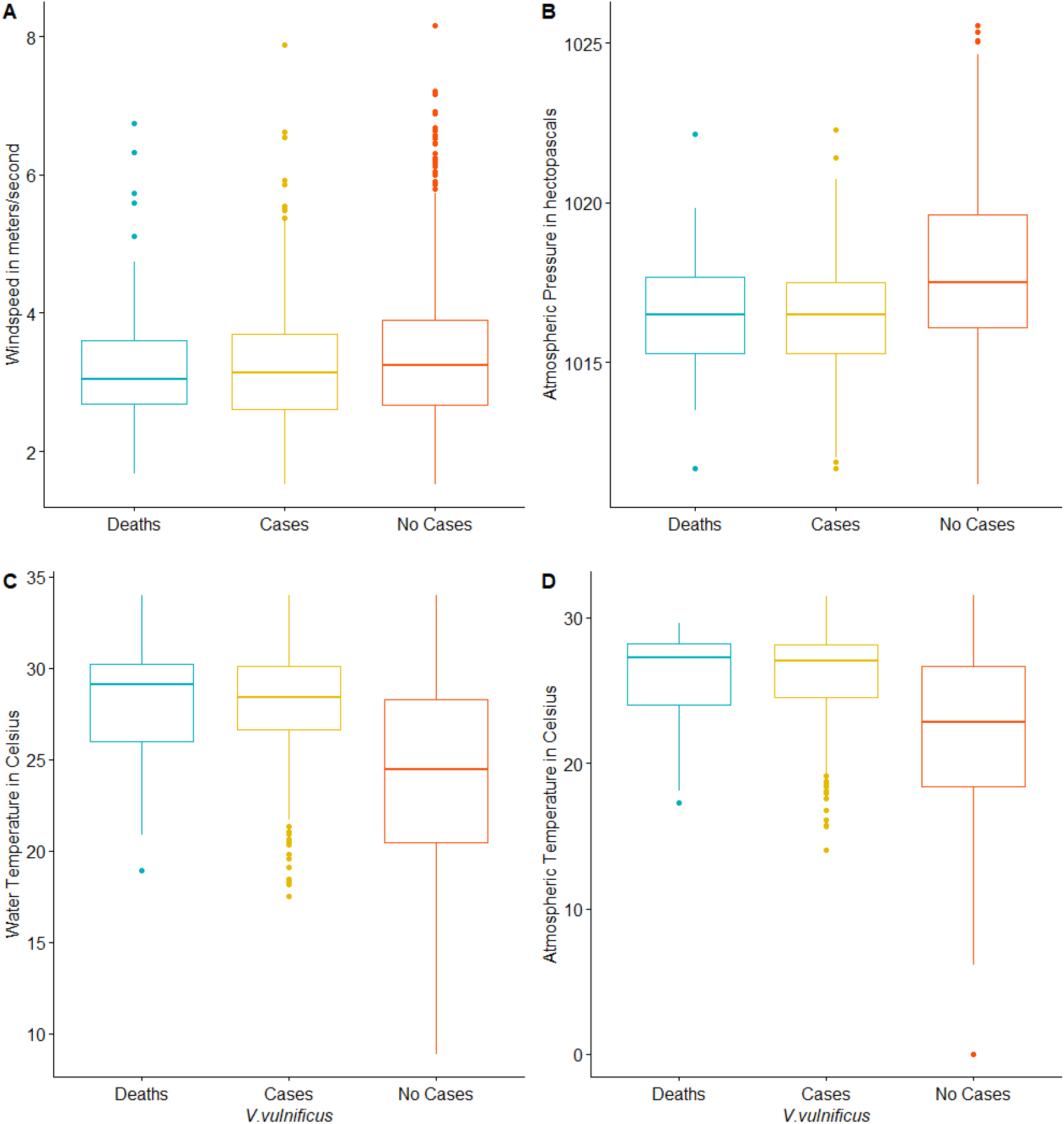
Correlations between various environmental variables and cases of V. vulnificus. A) wind speed (WSPD) in meters/second versus the incidence of *V. vulnificus* cases reported in Florida, stratified by *V. vulnificus* incidence, specifically during months when no cases were reported, months when cases with a survival outcome were reported, and months when mortality due to *V. vulnificus* infection was reported. The mean WSPD for months when no cases were reported was 3.35 ± 0.02 SE m/s, while the mean WSPD for months when cases where survival was the outcome was a mean of 3.26 ± 0.05 SE m/s, and the mean WSPD for months when death was the outcome was 3.27 ± 0.10 SE m/s. The Mann-Whitney U test indicated that the differences between the two groups were statistically significant (W = 325379, p-value = 0.01527). B) Atmospheric pressure (PRES) is stratified by *V. vulnificus* incidence according to months reporting deaths, months reporting cases, and months reporting no cases. The mean PRES for months when no cases were reported was 1017.8 ± 0.005 SE hPa while the mean PRES for months when cases where survival was the outcome was a mean of 1016.5 ± 0.003 SE hPa, and the mean PRES for months when death was the outcome was 1016.5 ± 0.002 SE hPa. C) Water temperature stratified by the reporting of *V. vulnificus*, specifically during months when no cases were reported, months when cases with a survival outcome were reported, and months when mortality due to *V. vulnificus* was reported. We found that our WTMP Kruskal-Wallis test was significant for this analysis, (*χ*^2^ = 176.14, df = 2, p-value < 0.0001), however, similar to ATMP, a pairwise multiple comparisons test with a Bonferroni adjustment demonstrated no significant difference between WTMP when survival cases were reported, and WTMP when non-survival cases were reported (p-value = 0.25). D) Atmospheric temperature (ATMP) stratified by *V. vulnificus* incidence, specifically during months when no cases were reported, months when cases with a survival outcome were reported, and months when mortality due to *V. vulnificus* was reported. As ATMP increases, so too does the likelihood of the severity of *V. vulnificus* cases. While this was a significant relationship (ATMP Kruskal-Wallis *χ*^2^ = 181.32, df = 2, p-value < 0.0001), a follow-up pairwise multiple comparison test with a Bonferroni adjustment demonstrated that the differences in ATMP between months when cases were reported and when cases were not reported was significant (p<0.0001), but not during months when survival cases were reported, and non-survival cases were reported (p =0.72). Please see main text for additional details.

#### b) Air Temperature (ATMP)

The mean ATMP for months when no cases were reported was 22.1 ± 1.15 SE °C, while the mean ATMP for months when cases where survival was the outcome was a mean of 25.9 ± 0.17 SE °C, and the mean WSPD for months when death was the outcome was 27.9 ± 0.36 SE °C (Fig. 3D). Using the Shapiro-Wilks test of Normality, we found that ATMP deviated from a Gaussian distribution for all stratifications (W = 0.94786, p-value < 0.0001) when no cases were reported, (W = 0.86938, p-value = <0.0001) when survival was reported, and (W = 0.83798, p-value = <0.0001) when death was reported.

Thus, given that mean ATMP was highest for death, followed by survival, and lastly for months when no cases were reported, we assessed the differences between the three groups using a non-parametric Kruskal-Wallis test of independent groups. While this was significant (Kruskal-Wallis *χ*^2^ = 181.32, df = 2, p-value < 0.0001), a follow-up pairwise multiple comparison test with a Bonferroni adjustment demonstrated that the differences in ATMP between ATMP during months when cases were reported and cases were not reported was significant (p<0.0001), but not during months when survival cases were reported, and non-survival cases were reported (p =0.72). Since this test was not significant, we pooled our case data to test the mean of the two independent groups, utilizing the Mann-Whitney U test to compare the means between the ATMP of the months when cases were reported, and months when cases were not reported. The Mann-Whitney U test indicated that the differences between the two groups were statistically significant (W = 508627, p-value = <0.0001), with the mean ATMP decreased during the months that no cases were reported, in contrast to months when cases were reported, regardless of their outcome. Overall, there appears to be a strong correlation between increased ambient air temperature and V. vulnificus infections that lead to patient death.

#### c) Water Temperature (WTMP)

The mean WTMP for months when no cases were reported was 24.5 ± 0.02 SE °C, while the mean WTMP for months when cases where survival was the outcome was a mean of 27.9 ± 0.17 SE °C, and the mean WTMP for months when death was the outcome was 28.2 ± 0.33 SE °C (Fig. 3C). To assess whether the distribution of each stratification deviated from the Gaussian distribution, we utilized the Shapiro-Wilks test of Normality. The data showed that WTMP had a non-normal distribution for all stratifications, e.g., (W = 0.96311, p-value < 0.0001) when no cases were reported, (W = 0.91159, p-value = <0.0001) when survival was reported, and (W = 0.92052, p-value = <0.0001) when death was reported.

Given that mean WTMP was highest for death, followed by survival, and lastly for months when no cases were reported, we assessed the differences between the three groups using a non-parametric Kruskal-Wallis test of independent groups. We found that our Kruskal-Wallis test was significant for this analysis, (Kruskal-Wallis *χ*^2^ = 176.14, df = 2, p-value < 0.0001), however, similar to ATMP, a pairwise multiple comparisons test with a Bonferroni adjustment demonstrated no significant difference between WTMP when survival cases were reported, and WTMP when non-survival cases were reported (p-value = 0.25). We followed up this analysis by pooling our observations for WTMP when cases were reported, regardless of outcome, in contrast to months when no cases were reported. A Mann-Whitney U test was performed, and this test was significant between the two independent groups (W = 451022, p-value < 0.0001). Overall, the data indicates that there appears to be a strong correlation between increased ambient air temperature and Vibrio vulnificus infections that lead to patient death

#### d) Pressure at Sea level (PRES)

The mean PRES for months when no cases were reported was 1017.8 ± 0.005 SE hPa while the mean PRES for months when cases where survival was the outcome was a mean of 1016.5 ± 0.003 SE hPa, and the mean PRES for months when death was the outcome was 1016.5 ± 0.002 SE hPa (Fig. 3B). We examined whether each stratification’s distribution deviated from the Gaussian distribution using the Shapiro-Wilks test of Normality. The data showed that PRES had a non-normal distribution for two of three stratifications, e.g., (W = 0.988, p-value < 0.0001) when no cases were reported, (W = 0.98942, p-value = 0.01721) when survival was reported, and (W = 0.98437, p-value = 0.4756) when death was reported. Given that mean PRES was extremely similar for months when cases survived, and when cases did not survive, we pooled our observations for PRES when cases were reported, regardless of the outcome, in contrast to months when no cases were reported. A Mann-Whitney U test was performed, and this test was significant between the two independent groups (W = 227799, p-value < 0.0001). In general, this suggests that as PRES decreases, the likelihood of *V. vulnificus* case reporting decreases.

### Combining the Environmental Factors into Single Models: Logistic Regression of Variables

We observed multiple significant correlations between two of our environmental variables and denoted that all four environmental variables had significant associations with *V. vulnificus* case reporting. Thus, we hypothesized that a statistical model composed of a combination of these variables would account for the variation in the reporting of clinical cases across the study sample. To do this, we used a logistic regression approach to compare different models.

Our logistic regression utilized AIC and McFadden’s R2 to identify the most parsimonious model that accounted for the variation in the likelihood of case occurrence seen in our data. The data suggested that the best model only accounted for 14.1% of the total variance. This model was generated by the independent variables WSPD + ATMP + WTMP + PRES + Month, with an AIC value of 1322.3. Month was a categorical variable with 12 levels, thus the months that generated a significant p-value were July (p-value = 0.015436), August (p-value = 0.013719), September (p-value = 0.0004), October (p-value = <0.0001), and November (p-value = 0.031814). In effect, although individual environmental factors were correlated with clinical cases, combinations of these factors into a single model only explained, at best, roughly 14% of the variation in case reporting.

## DISCUSSION

In 1999, Linkous and Oliver stated, “While *V. vulnificus* alone is responsible for 95% of all seafood-related deaths in the United States, it is still a mystery why more people do not develop this infection [11].” Balasubramanian et al. (2022) reported that biotic and abiotic factors generate selective pressures in ecosystems, which facilitate the emergence of pathogenic traits in *V. vulnificus* [51]. Thus, in the present study, we assessed the association between temporal and environmental variables such as wind speed (WSPD), ambient air temperature (ATMP), water temperature (WTMP), and pressure at sea level (PRES) in comparison to the reporting of *V. vulnificus* cases. It was not surprising that WTMP and ATMP were both significantly correlated with one another and demonstrated a strong relationship according to their *ρ* correlation value, regardless of whether cases were reported in a month. More surprising patterns involving WSPD: correlations were statistically significant with a higher *ρ* correlation value with ATMP and WTMP during months when cases and deaths were reported, as opposed to when no cases were reported. Given the negative values attributed to the *ρ* values, this suggests that as WSPD increased, WTMP and ATMP decreased during months when cases and deaths were reported. It was also interesting to note that WSPD and PRES were statistically significant during months when no cases were reported.

In addition, each of these was statistically significant when analyzed univariately, but none more than WTMP. PRES and WSPD, which were negatively correlated with *V. vulnificus* cases. These did not, however, account for explaining a notable amount of variation in the larger logistic regression model. Given that our environmental variables, coupled with the variable ‘Month’ account for only 14.1% of the variance of the likelihood between a case being reported versus not being reported, approximately 85% of the variance remain to be explained. This suggests these three variables are not sole predictors of *V. vulnificus* cases, and that the true cause of emergence likely contains many more drivers. This is consistent with modern understandings of disease emergence as being a complex system, composed of interactions between molecular and ecological factors [51, 52, 53, 54]. We speculate that epidemiological and microbial factors, such as host susceptibility and strain of *V. vulnificus*, may be more important to the development of an infection as opposed to the environmental variables that we measured. Other environmental variables not measured in this analysis, such as pH and salinity, may contribute to the explanatory variance in a logistic model [55, 56]. Indeed, these highly significant variables, including rainfall events, have been reviewed in-depth in the literature as they relate to *V. vulnificus* and related *Vibrio* species [22, 57, 58].

In our dataset, peak infections were reported in the late summer and fall, likely due to the water temperature around coastal Florida at that time. In our preliminary analysis of all 448 cases, *V. vulnificus* cases were most frequently reported in the months of July through September. When correcting for coastal cases that contained corresponding environmental variables, we found that mean WTMP was highest during those months when *V. vulnificus* was most frequently reported, e.g., July (30°C), August (30.1°C), and September (29.3°C). This data dovetails with the literature. In 1998, Motes et al. calculated the density of *V. vulnificus* in Atlantic coast oysters (*Crassostrea virginica*) from sites located on the Gulf coast in contrast to the Atlantic. *V. vulnificus* in oysters from the Gulf coast peaked between the months of May and October, which is similar to the frequency of human cases of *V. vulnificus* reported across the coastal cases of Florida in our dataset [14]. In areas of the globe where *V. vulnificus* has been detected or reported in water or shellfish, a correlation between water temperature and the density and/or abundance of *V. vulnificus* has been repeatedly demonstrated [19, 20, 24, 25, 50, 59, 60, 61, 62, 63, 64]. In our present study, we also show an association between water temperature and the incidence of *V. vulnificus* clinical cases. This however, begets the question, how will the incidence of *V. vulnificus* change over time as global climate change causes sea surface temperatures to continually increase [65]?

In Israel, Paz et al. (2007) examined the association between climate change, specifically warming temperatures, and the increased incidence of *V. vulnificus* in fish workers [59]. Since then, the topic of climate change and its relationship to *V. vulnificus* has garnered increasing attention [66, 67, 68, 69]. Deeb et al. reported that the incidence of all *Vibrio* infections has risen by 41% in the decade before 2005 [65]. In our dataset, we report a reporting increase of 333% between 2008 and 2017. Climate change is a crucial avenue of research, given that sea surface temperatures, tropical cyclone activity, and precipitation are all expected to increase in the Gulf of Mexico by the year 2100 [70].

While it is difficult to directly measure the impact of hurricanes on the density of a virulent species of bacteria using our monthly case report approach, we were able to indirectly measure the effect of extreme storm events using the measurements of sea-level pressure (PRES), which decreases during storm events [71, 72]. In our univariate analyses, we found that PRES was negatively correlated with the frequency of *V. vulnificus* cases, – however, its effect was dampened when used to build the larger logistical models. As the number of hurricane days during the summer and fall months increases in the state of Florida [48, 73, 74], we predict that this trend will be more apparent.

Even when indirectly measured, a trending correlation with climate change factors and the frequency with which *V. vulnificus* cases are reported in Florida is evident. For instance, Klontz et al. (1988) reported 62 cases in Florida between 1981 and 1987, for a mean of 8.8 cases per year [5]. In this study, 37.3 cases were reported on average per year, for a total of 448 cases between 2008 and 2020. That number jumped to 50 cases in 2017, the year that the highest frequency of cases was reported, with 22% of those cases reporting death as an outcome (11 deaths). For comparison, in 2013, the CDC reported that between 2002 and 2007, per year, 95 cases were reported with 85 hospitalizations and 35 deaths globally [12].

Within Florida, our data demonstrates that there are counties that disproportionately account for most of the clinical cases that were reported. The top five reporting counties were Hillsborough, Escambia, Pinellas, Brevard and Broward. The former three lie on the Gulf coast of Florida, Brevard lies on the Atlantic coast, and Broward in southwestern Florida. While we can report on the patterns, our dataset cannot account for these spatial differences. More research into the current dataset is needed to determine whether the strains that lie off the coast of these counties have the potential to be more virulent in human hosts, than in counties where fewer cases have been reported. Interestingly, a similar trend was detailed by López-Pérez et al. 2021, who reported differences in the virulence of *V. vulnificus* populations along the Atlantic coast of Florida [22]. Lastly, another reason for the rise in case reporting may be due to increased detectability via medical workers [7].

Finally, our case fatality rates were much lower than those reported in the literature [9, 75, 76] [9, 75, 76], for a mean of 24.78% across our 448-case dataset. Bross et al. reported case fatality rates of 50 percent for primary septicemia, and 15 percent for wound infections [77]. Klontz et al. noted a slightly higher trend for earlier wound infections in Florida, for a case fatality rate of 24% [5]. Given that our dataset did not distinguish between primary septicemia and wound infections, it is difficult to resolve why our case fatality rates were relatively low. One reason may be that medical personnel in Florida are now more attuned to the clinical signs of the disease than when it was initially characterized. Another may be that wound septicemia is the primary presentation, as opposed to the consumption of contaminated oysters, which carries a higher risk of mortality [15]. This supposed deviation should not surprise us, however, as *V. vulnificus* has already been demonstrated to have notable demographic and comorbidity factors that influence disease transmission [1, 2, 77]. And while our study identifies several key environmental actors that play a role in disease, further work is needed for truly holistic, multidimensional predictive models of *V. vulnificus* transmission and death.

## CONCLUSION

In this study, we have orchestrated a data science approach that combines reliable data from two different government sources—a public health (Florida Department of Health) and environmental (NOAA)—to establish correlations between environmental factors and clinical cases of disease caused by *V. vulnificus*. The findings reinforce the importance of the ecological dimension of disease emergence, where the infection process is just the final step in a multi-step process with multiple actors and forces. In particular, the relationship between environmental variables associated with elevated temperature and storm conditions is germane to conversations about how climate change may influence disease emergence and implore the development of more rigorous models that incorporate environmental actors in our effort to predict outbreaks of emerging diseases like *V. vulnificus*.

## Data Availability

All data produced in the present study are available upon reasonable request to the authors

## ACKNOWLEDGEMENTS

SAM was supported by a National Science Foundation CAREER award (#2045671). AJA was supported by a National Science Foundation Postdoctoral Fellowship in Biology (#2010904). The authors would also like to thank the Florida Department of Health for providing the data upon this work was performed. The authors would like to thank members of the Almagro-Moreno and Ogbunu research groups for helpful feedback.

## DATA AVAILABILITY

Interested parties are encouraged to contact the corresponding authors (AJA, SAM, and CBO) with regard to access to the data and code.

## AUTHOR CONTRIBUTIONS

AJA, SAM and CBO conceived the project. SAM and CBO supervised the project. AJA directed the data science pipeline. AJA, KM, CBO collected, curated, and analyzed the data. AJA, SAM and CBO interpreted and integrated the results. AJA, KM, SAM and CBO contribute to writing and editing the manuscript.

## REFERENCES

[1] Jani Anttila, Veijo Kaitala, Jouni Laakso, and Lasse Ruokolainen. Environmental variation generates environmental opportunist pathogen outbreaks. PLOS ONE, 10(12):e0145511, 2015.

[2] Annette Prüss, David Kay, Lorna Fewtrell, and Jamie Bartram. Estimating the burden of disease from water, sanitation, and hygiene at a global level. Environmental Health Perspectives, 110(5):537–542, 2002.

[3] Stuart Batterman, Joseph Eisenberg, Rebecca Hardin, Margaret E. Kruk, Maria Carmen Lemos, Anna M. Michalak, Bhramar Mukherjee, Elisha Renne, Howard Stein, Cristy Watkins, and Mark L. Wilson. Sustainable control of water-related infectious diseases: A review and proposal for interdisciplinary health-based systems research. Environmental Health Perspectives, 117(7):1023–1032, 2009.

[4] J J Farmer. The family Vibrionacea. In M Dworkin, S Falkow, E Rosenberg, K-H Schleifer, and E Stackebrandt, editors, The Prokaryotes: A Handbook on the Biology of Bacteria Volume 6: Proteobacteria: Gamma Subclas, pages 495–507. Springer New York, New York, NY, 2006.

[5] Karl C. Klontz. Syndromes of Vibrio vulnificus infections. Annals of Internal Medicine, 109(4):318, 1988.

[6] Shoukai Yu. Uncovering the geographical and host impacts on the classification of Vibrio vulnificus. Evolutionary Applications, 11(6):883–890, 2018.

[7] Po-Ren Hsueh, Ching-Yih Lin, Hung-Jen Tang, Hsin-Chun Lee, Jien-Wei Liu, Yung-Ching Liu, and Yin-Ching Chuang. Vibrio vulnificus in Taiwan. Emerging Infectious Diseases, 10(8):1363–1368, 2004.

[8] Craig Baker-Austin, Louise Stockley, Rachel Rangdale, and Jaime Martinez-Urtaza. Environmental occurrence and clinical impact of vibrio vulnificus and vibrio parahaemolyticus: a european perspective. Environmental Microbiology Reports, 2(1):7–18, 2010.

[9] WG Hlady, RC Mullen, and Hopkins RS. Vibrio vulnificus infections associated with raw oyster consumption -Florida, 1981-1992. Morbidity and Mortality Weekly Report, 42(21):405–450, 1993.

[10] Michael A. Horseman and Salim Surani. A comprehensive review of vibrio vulnificus: an important cause of severe sepsis and skin and soft-tissue infection. International Journal of Infectious Diseases, 15(3):e157–e166, 2011.

[11] Debra A Linkous and James D Oliver. Pathogenesis of Vibrio vulnificus. FEMS Microbiology Letters, 174(2):207–214, 1999.

[12] Sing-Peng Heng, Vengadesh Letchumanan, Chuan-Yan Deng, Nurul-Syakima Ab Mutalib, Tahir M. Khan, Lay-Hong Chuah, Kok-Gan Chan, Bey-Hing Goh, Priyia Pusparajah, and Learn-Han Lee. Vibrio vulnificus: An environmental and clinical burden. Frontiers in Microbiology, 8(997):1–14, May 2017.

[13] Melissa K. Jones and James D. Oliver. Vibrio vulnificus: Disease and pathogenesis. Infection and Immunity, 77(5):1723–1733, 2009.

[14] M. L. Motes, A. DePaola, D. W. Cook, J. E. Veazey, J. C. Hunsucker, W. E. Garthright, R. J. Blodgett, and S. J. Chirtel. Influence of water temperature and salinity on vibrio vulnificus in northern gulf and atlantic coast oysters (crassostrea virginica). Applied and Environmental Microbiology, 64(4):1459–1465, 1998.

[15] WG Hlady, RC Mullen, and RS Hopkin. Vibrio vulnificus from raw oysters. leading cause of reported deaths from foodborne illness in florida. The Journal of the Florida Medical Association, 80(8):536—538, 1993.

[16] Florida Department of Health. Vibrio vulnificus tallahassee, florida 2021. http://www.floridahealth.gov/diseases-and-conditions/vibrio-infections/vibrio-vulnificus/index.html, 2011. Online; accessed 12 December 2021.

[17] W. Gary Hlady. Vibrio infections associated with raw oyster consumption in florida, 1981–1994. Journal of Food Protection, 60(4):353–357, 1997.

[18] K. E. Weis, R. M. Hammond, R. Hutchinson, and C. G. M. Blackmore. Vibrio illness in florida, 1998–2007. Epidemiology and Infection, 139(4):591–598, 2010.

[19] Richard C. Tilton and Raymond W. Ryan. Clinical and ecological characteristics of vibrio vulnificus in the northeastern united states. Diagnostic Microbiology and Infectious Disease, 6(2):109–117, 1987.

[20] Jungsook Kim and Byung Chul Chun. Effect of seawater temperature increase on the occurrence of coastal vibrio vulnificus cases: Korean national surveillance data from 2003 to 2016. International Journal of Environmental Research and Public Health, 18(9):4439, 2021.

[21] Crystal N. Johnson, John C. Bowers, Kimberly J. Griffitt, Vanessa Molina, Rachel W. Clostio, Shaofeng Pei, Edward Laws, Rohinee N. Paranjpye, Mark S. Strom, Arlene Chen, Nur A. Hasan, Anwar Huq, Nicholas F. Noriea, D. Jay Grimes, and Rita R. Colwell. Ecology of vibrio parahaemolyticus and vibrio vulnificus in the coastal and estuarine waters of louisiana, maryland, mississippi, and washington (united states). Applied and Environmental Microbiology, 78(20):7249–7257, 2012.

[22] Mario López-Pérez, Jane M. Jayakumar, Trudy-Ann Grant, Asier Zaragoza-Solas, Pedro J. Cabello-Yeves, and Salvador Almagro-Moreno. Ecological diversification reveals routes of pathogen emergence in endemic vibrio vulnificus populations. Proceedings of the National Academy of Sciences, 118(40), 2021.

[23] A DePaola, G M Capers, and D Alexander. Densities of vibrio vulnificus in the intestines of fish from the u.s. gulf coast. Applied and Environmental Microbiology, 60(3):984–988, 1994.

[24] M. Tamplin, G. E. Rodrick, N. J. Blake, and T. Cuba. Isolation and characterization of vibrio vulnificus from two florida estuaries. Applied and Environmental Microbiology, 44(6):1466–1470, 1982.

[25] Mark A. Randa, Martin F. Polz, and Eelin Lim. Effects of temperature and salinity on vibrio vulnificus population dynamics as assessed by quantitative PCR. Applied and Environmental Microbiology, 70(9):5469–5476, 2004.

[26] DB Gilhousen. Improvements in national data buoy center measurements, 2007. National Data Buoy Center, Stennis Space Center, MS. 2007:4.

[27] NOAA. National Buoy Data Center Stennis Space Center, MS U.S. Department of Commerce. https://www.ndbc.noaa.gov/, 2021. Online; accessed 22 December 2021.

[28] National Data Buoy Center. Station ARPF1 - APK - Aripeka, FL Stennis Space Center, MS NOAA. https://www.ndbc.noaa.gov/station_page.php?station=trdf1, 2021.

[29] National Data Buoy Center. Station TRDF1 - 8721604 - Trident Pier, FL Stennis Space Center, MS: NOAA. https://www.ndbc.noaa.gov/station_page.php?station=arpf1, 2021.

[30] National Data Buoy Center. CLF1 - 8729840 - Pensacola, FL Stennis Space Center, MS: NOAA. https://www.ndbc.noaa.gov/station_realtime.php?station=pclf, 2021.

[31] Center NDB. Station 41114 - Fort Pierce, FL Stennis Space Center, MS: NOAA. https://www.ndbc.noaa.gov/station_page.php?station=41114, 2021.

[32] National Data Buoy Center. Station NPSF1 - 8725110 - Naples, FL Stennis Space Center, MS: NOAA. https://www.ndbc.noaa.gov/station_realtime.php?station=npsf1, 2021.

[33] National Data Buoy Center. Station LKWF1 - 8722670 - Lake Worth Pier, FL Stennis Space Center, MS: NOAA. https://www.ndbc.noaa.gov/station_page.php?station=lkwf1, 2021.

[34] National Data Buoy Center. Station SAPF1 - 8726520 - St. Petersburg, Tampa Bay, FL Stennis Space Center, MS: NOAA. https://www.ndbc.noaa.gov/station_page.php?station=sapf1, 2021.

[35] National Data Buoy Center. Station FRDF1 - 8720030 - Fernandina Beach, FL Stennis Space Center, MS: NOAA. https://www.ndbc.noaa.gov/station_page.php?station=frdf1, 2021.

[36] National Data Buoy Center. Station VAKF1 - 8723214 - Virginia Key, FL Stennis Space Center, MS: NOAA. https://www.ndbc.noaa.gov/station_page.php?station=vakf1, 2021.

[37] National Data Buoy Center. Station SAUF1 - St. Augustine, FL Stennis Space Center, MS: NOAA. https://www.ndbc.noaa.gov/station_page.php?station=sauf1, 2021.

[38] National Data Buoy Center. Station VENF1 - Venice, FL Stennis Space Center, MS: NOAA. https://www.ndbc.noaa.gov/station_page.php?station=venf1, 2021.

[39] National Data Buoy Center. Station PCBF1 - 8729210 - Panama City Beach, FL NOAA: Stennis Space Center, MS. https://www.ndbc.noaa.gov/station_page.php?station=pcbf1, 2021.

[40] National Data Buoy Center. Station APCF1 - 8728690 - Apalachicola, FL Stennis Space Center, MS: NOAA. https://www.ndbc.noaa.gov/station_page.php?station=apcf1, 2021.

[41] National Data Buoy Center. Station KYWF1 - 8724580 - Key West, FL Stennis Space Center, MS: NOAA. https://www.ndbc.noaa.gov/station_page.php?station=KYWF1, 2021.

[42] National Data Buoy Center. Station KTNF1 - Keaton Beach, FL Stennis Space Center, MS: NOAA. https://www.ndbc.noaa.gov/station_page.php?station=Ktnf1, 2021.

[43] National Data Buoy Center. Station CDRF1 - Cedar Key, FL Stennis Space Center, MS: NOAA. https://www.ndbc.noaa.gov/station_page.php?station=cdrf1, 2021.

[44] Jacqueline Rhoads. Post-hurricane katrina challenge: Vibrio vulnificus. Journal of the American Academy of Nurse Practitioners, 18(7):318–324, 2006.

[45] JJ Wetz, AD Blackwood, JS Fries, ZF Williams, and RT Noble. Trends in total vibrio spp. and vibrio vulnificus concentrations in the eutrophic neuse river estuary, north carolina, during storm events. Aquatic Microbial Ecology, 53:141–149, 2008.

[46] Olivia D. Nigro, Aixin Hou, Gayatri Vithanage, Roger S. Fujioka, and Grieg F. Steward. Temporal and spatial variability in culturable pathogenic vibrio spp. in lake pontchartrain, louisiana, following hurricanes katrina and rita. Applied and Environmental Microbiology, 77(15):5384–5393, 2011.

[47] M. P. Menon, P. A. Yu, M. Iwamoto, and J. Painter. Pre-existing medical conditions associated with vibrio vulnificus septicaemia. Epidemiology and Infection, 142(4):878–881, July 2013.

[48] Kristi S. Shaw, John M. Jacobs, and Byron C. Crump. Impact of hurricane irene on vibrio vulnificus and vibrio parahaemolyticus concentrations in surface water, sediment, and cultured oysters in the chesapeake bay, MD, USA. Frontiers in Microbiology, 5:1–11, 2014.

[49] Daniela Ceccarelli and Rita R. Colwell. Vibrio ecology, pathogenesis, and evolution. Frontiers in Microbiology, 5:1–3, 2014.

[50] Courtney S. Pfeffer, M. Frances Hite, and James D. Oliver. Ecology of vibrio vulnificus in estuarine waters of eastern north carolina. Applied and Environmental Microbiology, 69(6):3526–3531, 2003.

[51] Deepak Balasubramanian, Mario López-Pérez, Trudy-Ann Grant, C. Brandon Ogbunugafor, and Salvador Almagro-Moreno. Molecular mechanisms and drivers of pathogen emergence. Trends in Microbiology, 2022.

[52] Timothy Y. James, L. Felipe Toledo, Dennis Rödder, Domingos Silva Leite, Anat M. Belasen, Clarisse M. Betancourt-Román, Thomas S. Jenkinson, Claudio Soto-Azat, Carolina Lambertini, Ana V. Longo, Joice Ruggeri, James P. Collins, Patricia A. Burrowes, Karen R. Lips, Kelly R. Zamudio, and Joyce E. Longcore. Disentangling host, pathogen, and environmental determinants of a recently emerged wildlife disease: lessons from the first 15 years of amphibian chytridiomycosis research. Ecology and Evolution, 5(18):4079–4097, 2015.

[53] Pieter T. J. Johnson, Alan R. Townsend, Cory C. Cleveland, Patricia M. Glibert, Robert W. Howarth, Valerie J. McKenzie, Eliska Rejmankova, and Mary H. Ward. Linking environmental nutrient enrichment and disease emergence in humans and wildlife. Ecological Applications, 20(1):16–29, 2010.

[54] Sarah E. Kidd, Yat Chow, Sunny Mak, Paxton J. Bach, Huiming Chen, Adrian O. Hingston, James W. Kronstad, and Karen H. Bartlett. Characterization of environmental sources of the human and animal pathogen cryptococcus gattii in british columbia, canada, and the pacific northwest of the united states. Applied and Environmental Microbiology, 73(5):1433–1443, 2007.

[55] Eva Chase and Valerie J. Harwood. Comparison of the effects of environmental parameters on growth rates of vibrio vulnificus biotypes i, II, and III by culture and quantitative PCR analysis. Applied and Environmental Microbiology, 77(12):4200–4207, 2011.

[56] M.J. Hijarrubia, B. Lázaro, E. Suñén, and A. Fernández-Astorga. Survival ofVibrio vulnificusunder pH, salinity and temperature combined stress. Food Microbiology, 13(3):193–199, 1996.

[57] Mario López-Pérez, Jane M. Jayakumar, Jose M. Haro-Moreno, Asier Zaragoza-Solas, Geethika Reddi, Francisco Rodriguez-Valera, Orr H. Shapiro, Munirul Alam, and Salvador Almagro-Moreno. Evolutionary model of cluster divergence of the emergent marine pathogen vibrio vulnificus: From genotype to ecotype. mBio, 10(1), February 2019.

[58] Brian M. Schuster, Anna L. Tyzik, Rachel A. Donner, Megan J. Striplin, Salvador Almagro-Moreno, Stephen H. Jones, Vaughn S. Cooper, and Cheryl A. Whistler. Ecology and genetic structure of a northern temperate vibrio cholerae population related to toxigenic isolates. Applied and Environmental Microbiology, 77(21):7568–7575, 2011.

[59] S. Paz, N. Bisharat, E. Paz, O. Kidar, and D. Cohen. Climate change and the emergence of vibrio vulnificus disease in israel. Environmental Research, 103(3):390–396, 2007.

[60] L. Høi, J. L. Larsen, I. Dalsgaard, and A. Dalsgaard. Occurrence of vibrio vulnificus biotypes in danish marine environments. Applied and Environmental Microbiology, 64(1):7–13, 1998.

[61] R. L. Shapiro, S. Altekruse, L. Hutwagner, R. Bishop, R. Hammond, S. Wilson, B. Ray, S. Thompson, R. V. Tauxe, P. M. Griffin, and the Vibrio Working Group. The role of gulf coast oysters harvested in warmer months in vibrio vulnificus infections in the united states, 1988–1996. The Journal of Infectious Diseases, 178(3):752–759, 1998.

[62] J. Veenstra, P. J. G. M. Rietra, J. M. Coster, E. Slaats, and S. Dirks-Go. Seasonal variations in the occurrence of vibrio vulnificus along the dutch coast. Epidemiology and Infection, 112(2):285–290, 1994.

[63] Carla Hernández-Cabanyero, Eva Sanjuán, Belén Fouz, David Pajuelo, Eva Vallejos-Vidal, Felipe E. Reyes-López, and Carmen Amaro. The effect of the environmental temperature on the adaptation to host in the zoonotic pathogen vibrio vulnificus. Frontiers in Microbiology, 11:1–17, 2020.

[64] Yao Hsien Tey, Koa-Jen Jong, Shin-Yuan Fen, and Hin-Chung Wong. Occurrence of vibrio parahaemolyticus, vibrio cholerae, and vibrio vulnificus in the aquacultural environments of taiwan. Journal of Food Protection, 78(5):969–976, 2015.

[65] Reem Deeb, Daniel Tufford, Geoffrey I. Scott, Janet Gooch Moore, and Kirstin Dow. Impact of climate change on vibrio vulnificus abundance and exposure risk. Estuaries and Coasts, 41(8):2289–2303, 2018.

[66] Jaime Martinez-Urtaza, John C. Bowers, Joaquin Trinanes, and Angelo DePaola. Climate anomalies and the increasing risk of vibrio parahaemolyticus and vibrio vulnificus illnesses. Food Research International, 43(7):1780–1790, 2010.

[67] Brett A. Froelich and Dayle A. Daines. In hot water: effects of climate change on vibrio –human interactions. Environmental Microbiology, 22(10):4101–4111, 2020.

[68] Craig Baker-Austin, Joaquin Trinanes, Narjol Gonzalez-Escalona, and Jaime Martinez-Urtaza. Non-cholera vibrios: The microbial barometer of climate change. Trends in Microbiology, 25(1):76–84, 2017.

[69] Erin A. Urquhart, Benjamin F. Zaitchik, Darryn W. Waugh, Seth D. Guikema, and Carlos E. Del Castillo. Uncertainty in model predictions of vibrio vulnificus response to climate variability and change: A chesapeake bay case study. PLoS ONE, 9(5):e98256, 2014.

[70] Michela Biasutti, Adam H. Sobel, Suzana J. Camargo, and Timothy T. Creyts. Projected changes in the physical climate of the gulf coast and caribbean. Climatic Change, 112(3-4):819–845, 2011.

[71] Peter Dodge, Robert W. Burpee, and Frank D. Marks. The kinematic structure of a hurricane with sea level pressure less than 900 mb. Monthly Weather Review, 127(6):987–1004, 1999.

[72] Leo H. Holthuijsen, Mark D. Powell, and Julie D. Pietrzak. Wind and waves in extreme hurricanes. Journal of Geophysical Research: Oceans, 117(C9):987–1004, 2012.

[73] Michael E. Mann, Jonathan D. Woodruff, Jeffrey P. Donnelly, and Zhihua Zhang. Atlantic hurricanes and climate over the past 1, 500 years. Nature, 460(7257):880–883, 2009.

[74] Edward J. Phlips, Susan Badylak, Natalie G. Nelson, and Karl E. Havens. Hurricanes, el niño and harmful algal blooms in two sub-tropical florida estuaries: Direct and indirect impacts. Scientific Reports, 10(1), 2020.

[75] Craig Baker-Austin and James D. Oliver. Vibrio vulnificus: new insights into a deadly opportunistic pathogen. Environmental Microbiology, 20(2):423–430, 2017.

[76] J. D. Oliver. Wound infections caused by vibrio vulnificus and other marine bacteria. Epidemiology and Infection, 133(3):383–391, 2005.

[77] M H Bross, K Soch, R Morales, and R B Mitchell. Vibrio vulnificus infection: diagnosis and treatment. American Family Physician, 76(4):539–544, 2007.

